# The effect of COVID-19 and socioeconomic inequalities on emergency department accesses for psychiatric conditions

**DOI:** 10.1101/2025.04.25.25326419

**Authors:** Chiara Schepisi, Martina Ventura, Anteo Di Napoli, Massimiliano Aragona, Roberta Ciampichini, Valeria Fano, Christian Napoli, Martina Pacifici, Claudio Rosini, Caterina Silvestri, Fabio Voller, Alberto Zucchi, Alessio Petrelli

**Affiliations:** National Institute for Health, Migration and Poverty (INMP), Rome, Italy; Health Protection Agency (ATS), Bergamo, Italy; Local Health Unit Roma 2, Rome, Italy; Epidemiology and Cancer Registry Unit, Regina Elena National Cancer Institute, Rome, Italy; Tuscany Regional Health Agency (ARS), Florence, Italy

## Abstract

**Background:** COVID-19 produced a detrimental effect on mental health that could not be appropriately addressed due to a reduced accessibility of healthcare services, as patients with socioeconomic vulnerabilities faced the largest barriers in accessing healthcare during the pandemic. This study aimed to investigate emergency department (ED) accesses with a psychiatric diagnosis before and after COVID-19, with a specific focus on the role of socioeconomic factors.

**Methods:** Population-based longitudinal open cohort of residents and NHS beneficiaries aged ≥ 10, was enrolled in three large Italian areas from 1 January 2018 to 31 December 2021 (n=5,159,363). The primary outcome of interest was the first access to an ED (FMHEA); sex, age, deprivation level, citizenship and groups of diagnosis were considered as covariates. Crude incidence rates were calculated comparing the two time periods (pre- and post-COVID-19) by each covariate. In the context of interrupted time series (ITS), incidence rate ratios (IRR) of FMHEAs with 95%CI were estimated via an ITS analysis using a step-change negative binomial model. Temporal trend of FMHEAs was investigated through an ITS analysis on their monthly numbers.

**Results:** During follow-up, 56,762 FMHEAs were observed. The multivariate analysis showed that before COVID-19, the rate of FMHEA was higher among the highly deprived (IRR: 1.28; 95%CI:1.22.-1.35 for highest vs lowest deprivation level), females, people aged ≤24 and ≥85 and immigrants. A significant interaction between the COVID-19 period and deprivation level was found: the probability of reduced FMHEA increased with the deprivation level. The greater impact of COVID-19 was found in the central age classes resulting in a reduction in the differences in FMHEA rates by age in the post-Covid-19 period.

**Conclusion:** COVID-19 reduced incidence rates of ED accesses, but increased socioeconomic inequalities in the use of ED for mental health disorders.

## Introduction

Since the World Health Organization (WHO) declared the COVID-19 pandemic in March 2020 (WHO, 2020), countries worldwide implemented massive public health measures such as social distancing, isolation of infected patients and their contacts, and closure of non-urgent health services, schools, non-essential business, and gathering places. As a result, COVID-19 produced a profound socioeconomic crisis [1], particularly in those with pre-existing socioeconomic disadvantages [2,3]. The effects of the COVID-19 pandemic and its socioeconomic sequelae on mental health have been extensively explored, with findings suggesting that adverse, though short lasting, consequences may develop [4,5,6,7], especially in vulnerable subsets of the population [8,9,10].

While the mental health burden among the population increased, delivery of mental health care was challenged by workforce shortages and burnout in mental healthcare providers related to increased workload and safety concerns [11]. Therefore, the number of psychiatric visits decreased during the first wave of COVID-19 across European countries [12]. In Italy, 13% of Community Mental Health Services closed, and 25% reduced their opening hours during the early pandemic phase.

Moreover, most day hospitals, which provide close clinical monitoring to subacute patients, were closed when lockdown was declared. As a result, all activities except urgent consultations and compulsory treatments were significantly limited [13]. Reduced access to healthcare is proven to have been unequal as it penalized the most deprived individuals and migrants [14, 15]). Unlike community and inpatient services, emergency departments (ED) remained available throughout the pandemic period, with no significant restriction. Therefore, investigating accesses to EDs for mental health conditions could provide useful insight into mental health needs throughout the COVID-19 pandemic phases. In Italy, few studies have investigated ED use for mental health complaints, although a decrease in ED visits for psychiatric conditions during the first wave has been shown [16], as has an increase in the incidence of ED accesses for any medical condition [17]. However, large population-based studies assessing the effect of COVID-19 on ED accesses and the role of socioeconomic inequalities are lacking.

This study aimed to investigate differences between pre- and post-COVID-19 in the use of EDs for psychiatric disorders, with a specific focus on the role of socioeconomic factors.

## Methods

This study is part of the larger COVID-19 and Mental Health (CoMeH) project, a multicentre collaborative project aimed at evaluating the impact of the COVID-19 pandemic on the use of mental health facilities in Italy, with a specific focus on the role of socioeconomic and citizenship status.

The CoMeH study has been promoted by the National Epidemiologic Observatory for Equity in Health (OENES) of the National Institute for Health, Migration and Poverty (INMP) and involves the Tuscany Regional Health Agency (RHA), the Bergamo Local Health Authority (LHA), and the Rome 2 LHA. These three centres cover nearly 10% (almost 6 million people) of Italian National Health Service (NHS) beneficiaries.

For the aim of the CoMeH project, we enrolled an open cohort of subjects starting from 01 January 2018, with enrolment until the end of December 2024. Data in the present study are updated to 31 December 2021. The cohort was created retrospectively from the Municipal Registries and the General Practitioner (GP) Registries of each participating centre. Inclusion criteria were age ≥10 years, having been a resident for at least two years before enrolment in any of the centres’ catchment areas, and being assisted by an NHS GP.

Cohort exit criteria were (i) end of the planned period (31 December 2021) (ii) death, or (iii) emigration, whichever occurred first. Given the open cohort design, individuals who exited the cohort for emigration could be re-admitted if they resettled in the health centres’ catchment areas. A more detailed description of the study protocol has been published previously [18].

### Outcomes

The main outcomes of the study were the incidence of the first ED access of individuals with a psychiatric condition (First Mental Health Emergency Access–FMHEA), either as a primary diagnosis or associated with any other medical condition. We also included the secondary diagnosis, as some psychiatric acute episodes may lead to severe medical consequences which could be the main reason for urgent access (i.e., a patient with a depressive episode with a first diagnosis of traumatic injury due to suicide attempt). Psychiatric diagnosis was based on the International Classification of Disease, 9^th^ Revision, Clinical Modification (ICD-9-CM).

Specifically, we grouped the psychiatric diagnoses into 15 clinical categories, as listed in Table 1. Data on FMHEAs were retrieved through the emergency care information system. Only incident accesses, identified as those events without any other access for any of the above-mentioned ICD-9 CM codes in the two years prior to the event date, were considered as outcomes.

**Table 1.** Classification of psychiatric conditions based on ICD9 CM codes of diagnosis.

| Group of diagnoses | ICD9 CM Codes |
| --- | --- |
| Delirium/Mental Confusion | 291.0, 292.81, 293.0, 293.1, 298.2 |
| Dependencies | 291-292, 303-305 |
| Dissociation | 300.10-300.15, 300.6 |
| Eating Disorders | 307.1, 307.5, 307.50, 307.51 |
| Anxiety Disorders | 293.84, 293.89, 300.00-300.02, 300.09, 300.20-300.23, 300.29, 300.3, 300.5 |
| Sleep Disorders | 307,4 |
| Mood Disorders | 293.83, 296, 298.0, 300.4, 309.1, 311 |
| Personality Disorders | 301 |
| Schizoaffective Disorder | 295,7 |
| Unspecified Psychoses | 293.81, 293.82, 298, 298.1, 298.4, 298.8, 298.9, 299.90, 299.91 |
| Adjustment Disorders | 309 (except 309.81) |
| Post-Traumatic Stress and related Disorders | 308, 309.81 |
| Somatization | 300.7, 300.8, 307.8 |
| Schizophrenic Spectrum | 295, 295.0-295.6, 295.8-295.9, 297.0-297.9, 298.3 |

### Covariates

The sociodemographic information of the individuals enrolled in the cohort was retrieved from the municipal population registers. Age, sex, citizenship, municipality, and deprivation index were considered as covariates. Subjects were classified into six classes based on their age at the time of FMHEA: <24, 25–34, 35–64, 65–74, 75–84, and >85 years. Differences in socioeconomic status were investigated using the national deprivation index, an area-based index which is calculated for each census tract as the sum of five standardized indicators and categorized into quintiles: low education level (% of the population with elementary school education or less); unemployment (% of the working age population that is unemployed or searching for first job); no ownership of dwelling (% of dwellings that are rented); single-parent families (% of single-parent families with minor children making up one household); residential occupant density (population per 100 square meters). A deprivation level was attributed to each subject through a record linkage with the census tract of residence. As census tracts are very small areas, the deprivation index can be considered a good proxy of individual socioeconomic status.

In order to investigate the impact of the pandemic on the immigrant population, the cohort was stratified based on citizenship into Italians and immigrants. All the residents in Italy without Italian citizenship were considered as immigrants. In Italy, citizenship rather than country of birth is considered the best proxy of immigrant status, at least when assessing the most recent immigrations [19]; in fact, immigrants can obtain Italian citizenship only either by marriage or by application after from three to 10 consecutive years of legal residence. Moreover, children born in Italy to foreign parents can obtain citizenship after their 18^th^ birthday. Immigrants from highly developed countries (HDCs) were included in the Italians group as they accounted for only about 5% of the foreign resident population in the cohort and generally have a health profile comparable to that of the native population; immigrants from high migratory pressure countries (HMPCs) made up the immigrants group [20].

The study period was divided in two phases: pre-COVID-19 (from January 2018 to February 2020) and post-COVID-19 (from March 2020, when the WHO declared the COVID-19 pandemic, to December 2021).

### Ethics and Privacy

The study protocol was approved by the ethics committee of the Italian National Institute of Health (Istituto Superiore di Sanità; protocol n. 0029105, July 25, 2022). Authorization for the use of anonymized data was obtained from the Data Protection Officers of each participating centre according to EU regulation 2016/679.

Privacy protection was ensured by assigning each individual a validated anonymous patient identifier so that multiple data sources could be linked, thereby making it possible to follow up enrolled subjects. Any personally identifiable information was hidden from individual records.

### Statistical analysis

Baseline demographic and socioeconomic characteristics of the CoMeH study cohort and of the FMHEAs identified during the follow-up are described as frequencies and percentages.

To assess the effect of the pandemic on FMHEAs, crude incidence rates per 100,000 person-days with 95%CI were calculated overall and comparing the two time periods (pre-COVID-19 and post-COVID-19) by sex, age class, deprivation level, citizenship, and diagnosis.

In order to identify the impact of the COVID-19 outbreak on FMHEAs, we conducted two types of analysis, both in the context of interrupted time series (ITS).

In the first analysis, incidence rate ratios (IRR) of FMHEAs with 95%CI were estimated via an ITS analysis using a step-change negative binomial model. The model was built taking into account the presence of over-dispersion, with the log person-time of follow-up for each group as offset and adjusted for age class, sex, a binary period term (pre-pandemic and pandemic), citizenship, deprivation level, a linear time variable to account for trends, and a categorical calendar month variable to identify any seasonal effect. Moreover, to assess the effect of citizenship and deprivation level on the FMHEA trend before and after the pandemic outbreak, two interaction terms–between citizenship and time period and between deprivation level and time period–were included. The interaction between age and time period was also tested.

In the second analysis, the impact of the pandemic on the temporal trend of FMHEAs was investigated, estimating IRR, overall and stratified by sex, age class, citizenship, deprivation level, and diagnosis. Only diagnoses accounting for at least 150 ED accesses were analysed. The ITS analysis was conducted on the monthly number of FMHEAs. According to the procedure proposed by Schuengel et al. [21], data were detrended using Loess regression and smoothing, and seasonality was tested. If the presence of seasonal effects was identified, an adjustment was performed. The change in slope from the pre-pandemic to the pandemic period was tested using Poisson segmented regression with Newey-West standard errors to account for autocorrelation and heteroscedasticity. The pre-COVID-19 trend, the immediate effect of the outbreak of the pandemic, and the post-COVID-19 trend were evaluated. All statistical analyses were performed using SAS 9.3 and R Studio (version 4.1.3).

## Results

Baseline characteristics of the study population and of FMHEAs are described in Table 2. Overall, 5,159,363 subjects were enrolled in the cohort between 1 January 2018 and 31 December 2021. During follow-up, 56,762 FMHEAs were observed (1.1% of the population).

**Table 2.** Baseline characteristics of the study population and of first mental health emergency access.

| Baseline characteristics | Population |  | First MH Emergency Access |  |
| --- | --- | --- | --- | --- |
|  | N | % | N | % |
| **Total** | 5,159,363 |  | 56,762 | 1.10% |
| **Age class** |  |  |  |  |
| $\leq 24$ | 689,332 | 13.4% | 11,061 | 19.5% |
| 25-34 | 564,958 | 11.0% | 7,033 | 12.4% |
| 35-64 | 2,510,617 | 48.7% | 23,700 | 41.8% |
| 65-74 | 664,706 | 12.9% | 5,530 | 9.7% |
| 75-84 | 506,405 | 9.8% | 6,446 | 11.4% |
| 85+ | 223,345 | 4.3% | 2,992 | 5.3% |
| **Sex** |  |  |  |  |
| Males | 2,466,975 | 47.8% | 23,643 | 41.7% |
| Females | 2,692,388 | 52.2% | 33,119 | 58.3% |
| **Deprivation level (quintiles)** |  |  |  |  |
| Low | 1,026,856 | 19.9% | 10,442 | 18.4% |
| Middle-low | 1,193,167 | 23.1% | 12,839 | 22.6% |
| Middle | 1,065,511 | 20.7% | 12,080 | 21.3% |
| Middle- high | 753,674 | 14.6% | 8,582 | 15.1% |
| High | 464,521 | 9.0% | 5,888 | 10.4% |
| Missing data | 655,634 | 12.7% | 6,931 | 12.2% |
| **Citizenship** |  |  |  |  |
| Italians | 4,614,768 | 89.4% | 50,225 | 88.5% |
| Immigrants from HDCs | 22,650 | 0.4% | 199 | 0.4% |
| Immigrants from HPMCs | 446,334 | 8.7% | 5,661 | 10.0% |
| Missing data | 75,611 | 1.5% | 677 | 1.2% |
| **Group of diagnoses** |  |  |  |  |
| Delirium/Mental Confusion |  |  | 6,038 | 10.6% |
| Dependencies |  |  | 3,625 | 6.4% |
| Dissociation |  |  | 181 | 0.3% |
| Eating Disorders |  |  | 161 | 0.3% |
| Anxiety Disorders |  |  | 24,968 | 44.0% |
| Sleep Disorders |  |  | 52 | 0.1% |
| Mood Disorders |  |  | 4,586 | 8.1% |
| Personality Disorders |  |  | 441 | 0.8% |
| Schizoaffective Disorder |  |  | 7 | 0.0% |
| Unspecified Psychoses |  |  | 4,906 | 8.6% |
| Adjustment Disorders |  |  | 682 | 1.2% |
| Post-Traumatic Stress and<br>related Disorders |  |  | 1,219 | 2.1% |
| Somatization |  |  | 6,939 | 12.2% |
| Schizophrenic Spectrum |  |  | 279 | 0.5% |
| Others |  |  | 2,678 | 4.7% |

Compared to the general population, the patients accessing the ED with a psychiatric diagnosis were more frequently the youngest (19.5% vs 13.4% aged ≤24 years) and females (58.3% vs 52.2%), while the distributions by deprivation level and citizenship were similar.

Anxiety Disorders were the most frequent diagnoses (44%), followed by Somatization (12.2%), Delirium/Mental Confusion (10.6%), Unspecified Psychoses (8.6%), and Mood Disorders (8.1%).

Table 3 reports crude incidence rates of FMHEAs in the periods pre- and post-COVID-19, overall and by sex, age, deprivation level, citizenship, and group of diagnoses. Rates of FMHEAs decreased after the outbreak of the pandemic, overall and for all subgroups of the population observed, although at different strengths in the different groups. In the pre-pandemic period, higher rates were observed as the deprivation level increased, while during the pandemic, subjects living in areas with a high deprivation level experienced the highest reduction in rates. The youngest and the oldest patients had the highest rates of FMHEAs compared to other age classes in both periods, but the decrease after the outbreak of the pandemic was stronger in the central age classes. Immigrants from HMPCs had higher rates compared to Italians in the pre-pandemic period, but both groups experienced a similar reduction in rates during the pandemic. Analysing rates by group of diagnoses, a decrease was observed for all groups, with the only exception being Eating Disorders (albeit with a low number of accesses), which increased after the outbreak of the pandemic.

**Table 3.**
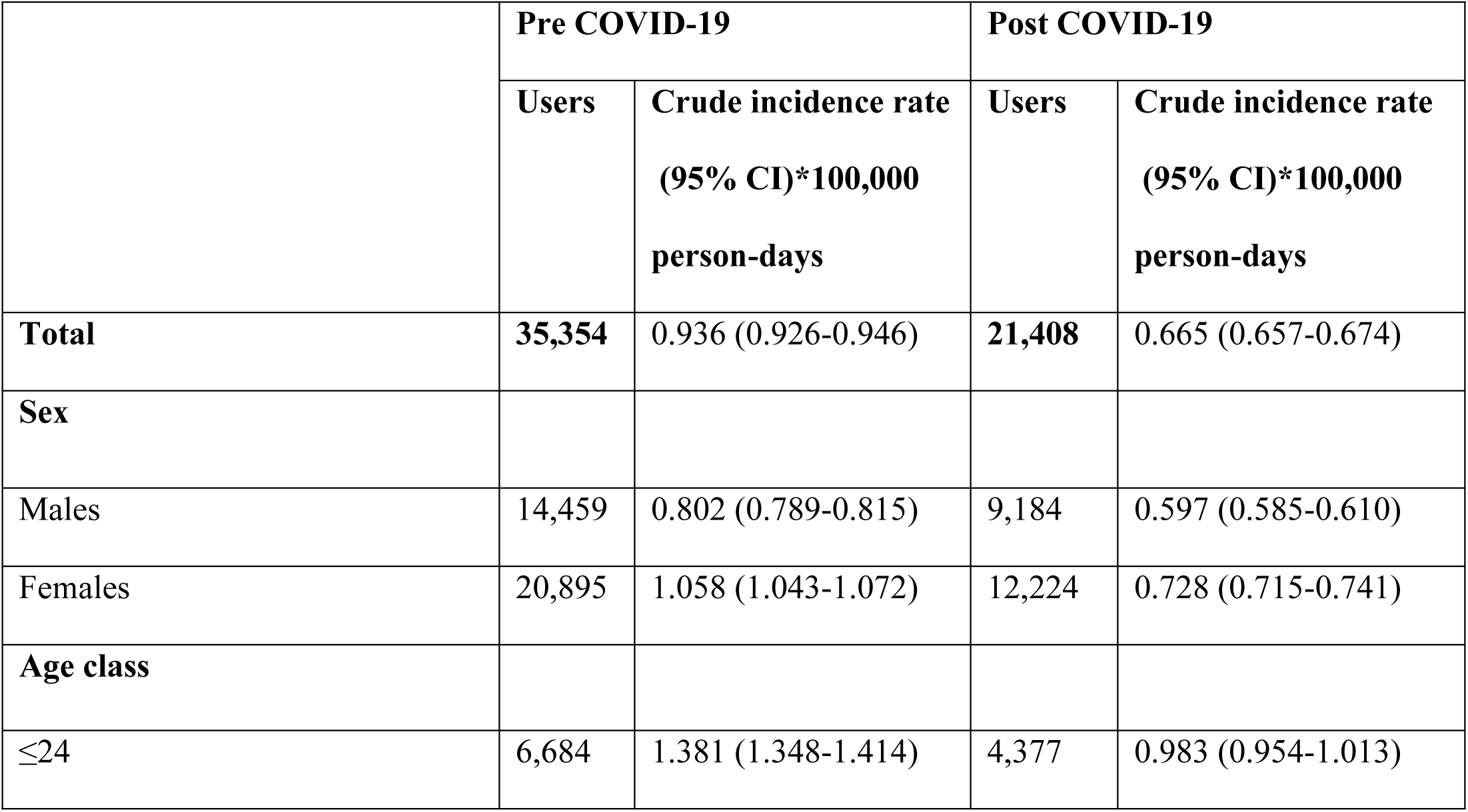
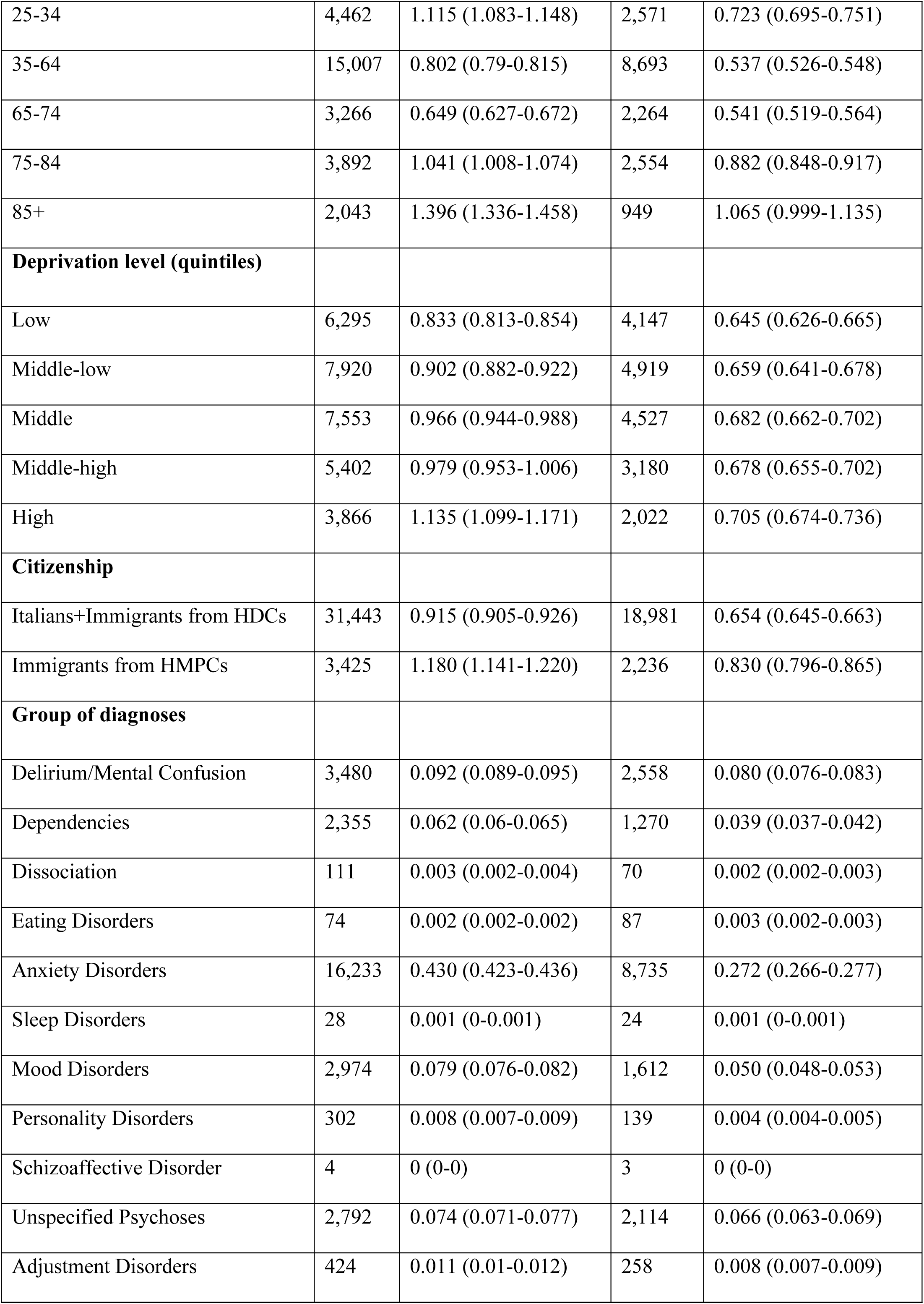
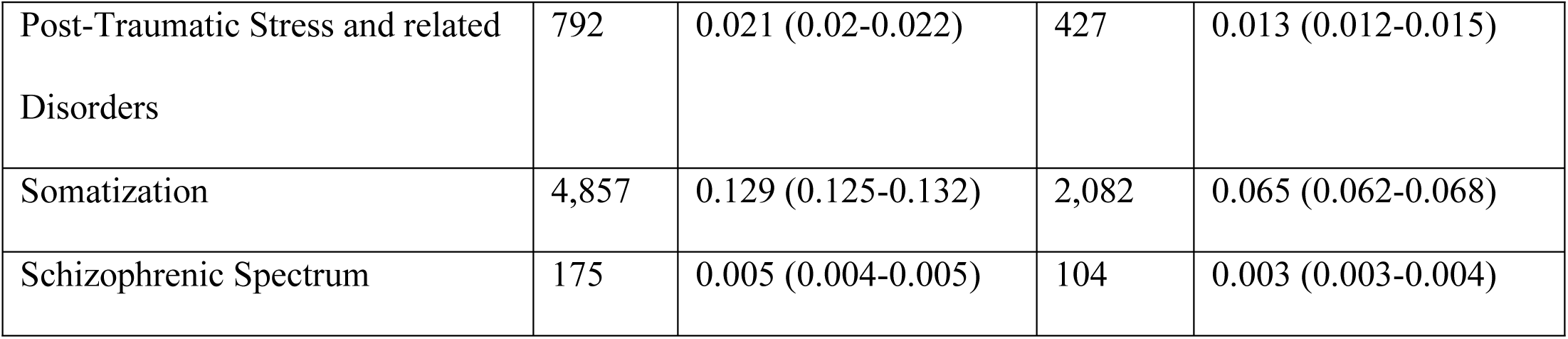
FMHEAs: Crude incidence rates *100,000 person-days by sociodemographic characteristics and group of diagnoses in pre- and post-COVID-19 periods.

The results of the multivariate ITS analysis are shown in Table 4. Model 1 included two interaction terms, respectively between citizenship and time period and between deprivation level and time period; in Model 2, instead, the interaction between age and time period was tested. After adjusting for age, sex, deprivation level, and citizenship, both models confirmed a significant reduction in FMHEAs in the pandemic period compared to the pre-pandemic one (Model 1: IRR=0.74 p<0.001; Model 2: IRR=0.69 p<0.001). The probability of having an FMHEA increased by deprivation level in the pre-pandemic period. A significant interaction between the COVID-19 period and deprivation level was also observed (p of interaction term <0.001): the probability of reduced FMHEA increased with the deprivation level (Figure 1). Instead, a higher adjusted rate was observed for immigrants from HMPCs compared to Italians. All age classes except the oldest showed lower probabilities of FMHEAs compared to the youngest class (≤ 24 years). When we tested the interaction between age and the COVID-19 period (Model 2), a significant effect modification was found, with a greater impact of COVID-19 in the central age classes (p interaction term <0.001): the effect resulted in a reduction in the differences in FMHEA rates by age in the post-Covid-19 period (Model 2, Fig 1).

**Fig. 1.**
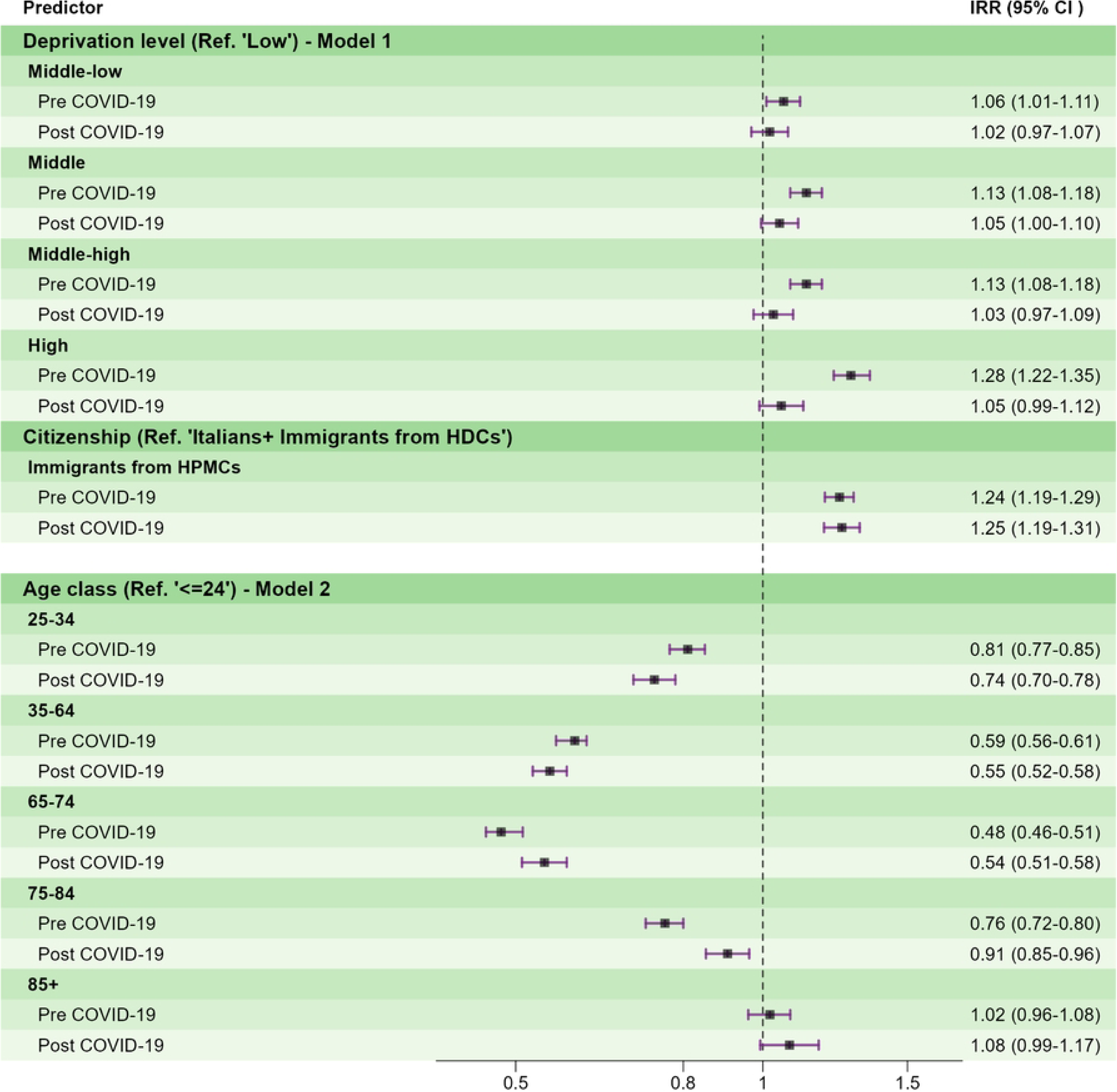
Forest plot of the multivariate ITS analysis. Model 1: interaction between deprivation and time period and between citizenship and time period. Model 2: interaction between age and time period. Pre-COVID-19 refers to the coefficient of each level of predictor. Post-COVID-19 refers to the sum of the coefficient of each level of predictor and the interaction term between the predictor and COVID-19.

**Table 4.** Results of ITS multivariate regression models. Adjusted incidence rate ratios and 95%CI of FMHEAs.

|  | Model1 |  |  |  | Model2 |  |  |
| --- | --- | --- | --- | --- | --- | --- | --- |
| Predictors | *IRR* | *CI* | *p* |  | *IRR* | *CI* | *p* |
| **Time period (ref. Pre-COVID-19)** |  |  |  |  |  |  |  |
| Post-COVID-19 | **0.74** | **0.70 – 0.79** | **<0.001** |  | **0.69** | **0.65 – 0.73** | **<0.001** |
| Time (continuous) | 1.00 | 1.00 – 1.00 | 0.173 |  | 1 | 1.00 – 1.00 | 0.153 |
| **Study Month (ref. January)** |  |  |  |  |  |  |  |
| February | 0.97 | 0.92 – 1.02 | 0.236 |  | 0.97 | 0.92 – 1.02 | 0.235 |
| March | **0.91** | **0.86 – 0.96** | **0.001** |  | **0.91** | **0.86 – 0.96** | **0.001** |
| April | **0.94** | **0.89 – 0.99** | **0.020** |  | **0.94** | **0.89 – 0.99** | **0.019** |
| May | 1.05 | 0.99 – 1.10 | 0.101 |  | 1.05 | 0.99 – 1.10 | 0.097 |
| June | **1.24** | **1.18 – 1.31** | **<0.001** |  | **1.24** | **1.18 – 1.31** | **<0.001** |
| July | **1.24** | **1.18 – 1.31** | **<0.001** |  | **1.24** | **1.18 – 1.31** | **<0.001** |
| August | **1.19** | **1.13 – 1.25** | **<0.001** |  | **1.19** | **1.13 – 1.25** | **<0.001** |
| September | **1.09** | **1.03 – 1.15** | **0.002** |  | **1.09** | **1.03 – 1.15** | **0.002** |
| October | 0.99 | 0.94 – 1.04 | 0.684 |  | 0.99 | 0.94 – 1.04 | 0.679 |
| November | 0.96 | 0.91 – 1.02 | 0.17 |  | 0.96 | 0.91 – 1.02 | 0.166 |
| December | 0.95 | 0.90 – 1.01 | 0.083 |  | 0.95 | 0.90 – 1.01 | 0.083 |
| **Deprivation Level (ref. Low)** |  |  |  |  |  |  |  |
| Middle-low | **1.06** | **1.01 – 1.11** | **0.009** |  | **1.04** | **1.01 – 1.08** | **0.014** |
| Middle | **1.13** | **1.08 – 1.18** | **<0.001** |  | **1.10** | **1.06 – 1.13** | **<0.001** |
| Middle-high | **1.13** | **1.08 – 1.18** | **<0.001** |  | **1.09** | **1.05 – 1.13** | **<0.001** |
| High | **1.28** | **1.22 – 1.35** | **<0.001** |  | **1.19** | **1.14 – 1.24** | **<0.001** |
| Citizenship (ref. Italians+HDCs) |  |  |  |  |  |  |  |
| Immigrants from HPMCs | 1.24 | 1.19 – 1.29 | <0.001 |  | 1.24 | 1.20 – 1.28 | <0.001 |
| Sex (ref. Females) |  |  |  |  |  |  |  |
| Males | 0.79 | 0.78 – 0.81 | <0.001 |  | 0.98 | 0.94 – 1.02 | 0.404 |
| Age class (ref. ≤24) |  |  |  |  |  |  |  |
| 25-34 | 0.78 | 0.75 – 0.81 | <0.001 |  | 0.81 | 0.77 – 0.85 | <0.001 |
| 35-64 | 0.57 | 0.55 – 0.59 | <0.001 |  | 0.59 | 0.56 – 0.61 | <0.001 |
| 65-74 | 0.50 | 0.48 – 0.53 | <0.001 |  | 0.48 | 0.46 – 0.51 | <0.001 |
| 75-84 | 0.81 | 0.78 – 0.85 | <0.001 |  | 0.76 | 0.72 – 0.80 | <0.001 |
| 85+ | 1.04 | 0.99 – 1.09 | 0.128 |  | 1.02 | 0.96 – 1.08 | 0.546 |
| Deprivation Level*Time period |  |  | p<0.001 |  |  |  |  |
| Middle-low | 0.96 | 0.90 – 1.03 | 0.261 |  |  |  |  |
| Middle | 0.93 | 0.87 – 0.99 | 0.025 |  |  |  |  |
| Middle-high | 0.91 | 0.85 – 0.98 | 0.013 |  |  |  |  |
| High | 0.82 | 0.76 – 0.89 | <0.001 |  |  |  |  |
| Citizenship *Time period |  |  | p=0.9 |  |  |  |  |
| Immigrants from HPMCs | 1.01 | 0.94 – 1.07 | 0.878 |  |  |  |  |
| Age group *Time period |  |  |  |  |  |  | p<0.001 |
| 25-34 |  |  |  |  | 0.91 | 0.85 – 0.99 | 0.021 |
| 35-64 |  |  |  |  | 0.94 | 0.88 – 1.00 | 0.043 |
| 65-74 |  |  |  |  | 1.13 | 1.04 – 1.22 | 0.004 |
| 75-84 |  |  |  |  | 1.19 | 1.10 – 1.29 | <0.001 |
| 85+ |  |  |  |  | 1.06 | 0.95 – 1.17 | 0.279 |

The results of the ITS analysis conducted on the trend of the monthly number of FMHEAs are shown in Table 5, Fig 2-3, and Supplementary Fig 1-5, overall and stratified by sex, age class, deprivation level, citizenship, and group of diagnoses. Overall, the average number of monthly FMHEAs was 1182.5 (SD=269.2). At the COVID-19 outbreak, overall FMHEAs significantly dropped (IRR: 0.636, 95%CI 0.481-0.840). In the post-COVID period, a moderate but significant increasing trend (IRR=1.018, 95%CI 1.001-1.036) was observed, which exceeded the counterfactual line (Fig 2).

**Fig. 2:**
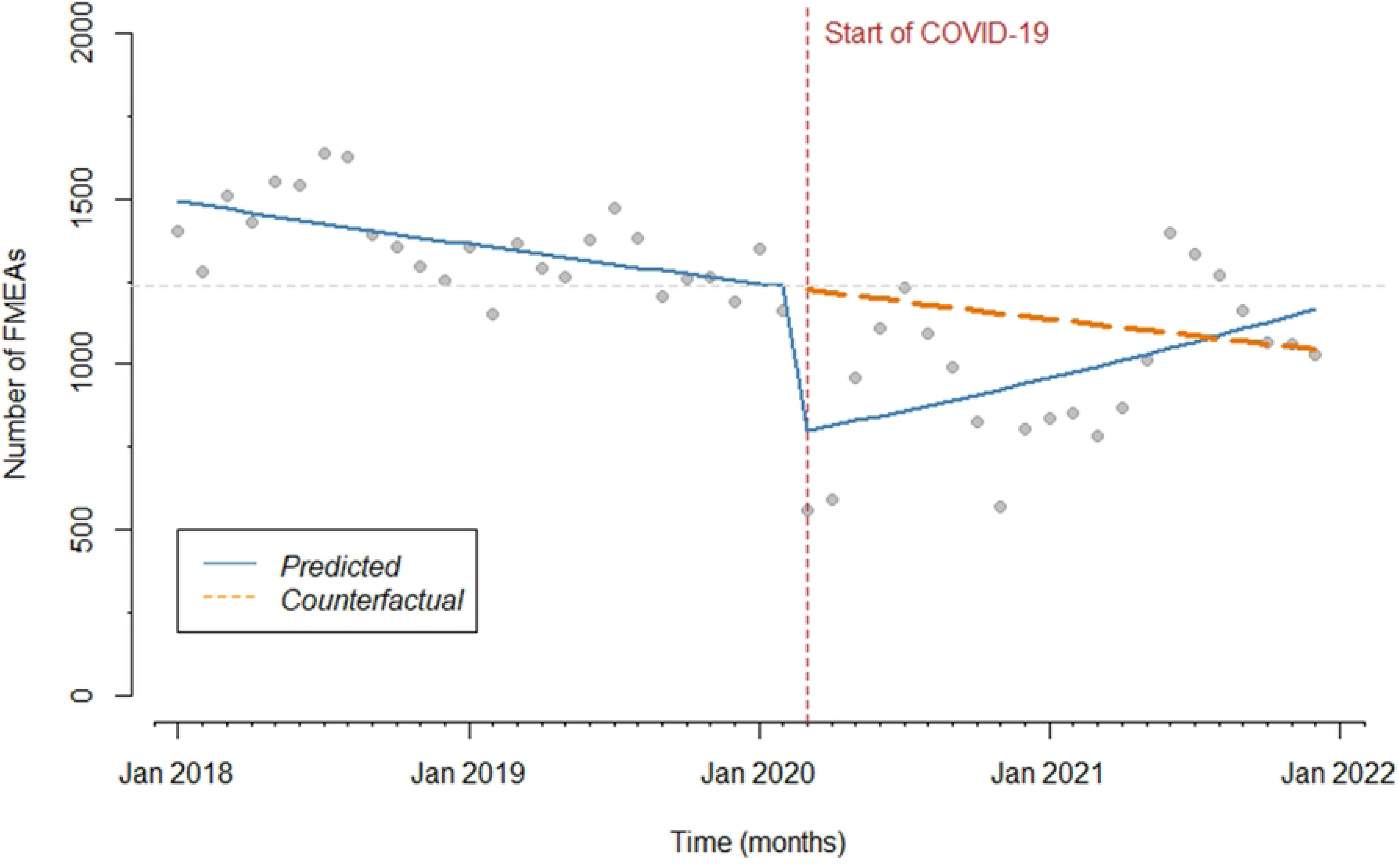
Number of monthly FMHEAs before and during the COVID-19 pandemic. Vertical dashed line: introduction of restrictions. Continuous line: trend over the years. Dashed line: counterfactual scenario. Horizontal dashed line: pre-pandemic level.

**Fig. 3:**
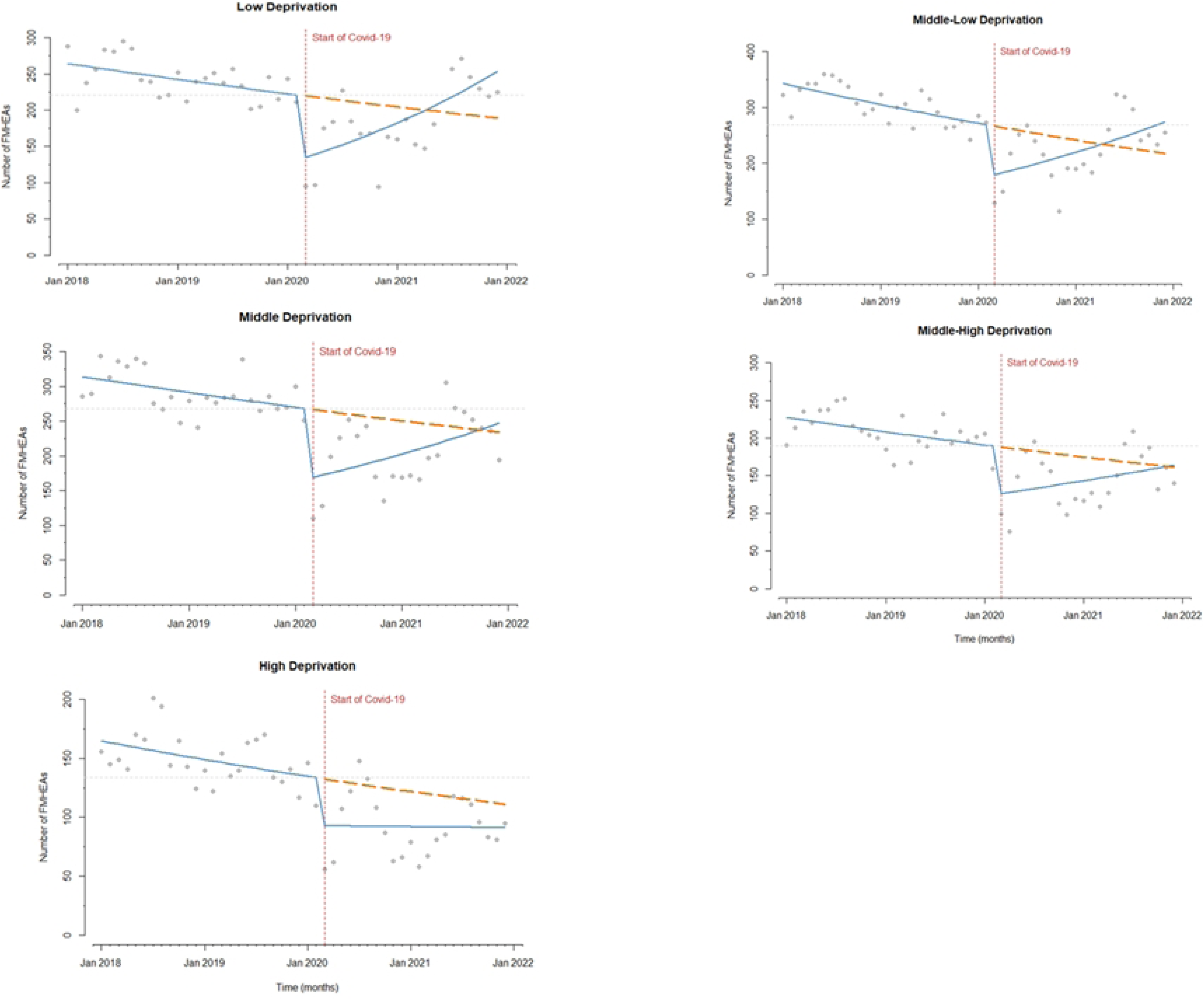
Number of monthly FMHEAs before and during the COVID-19 pandemic by deprivation level. Vertical dashed line: introduction of restrictions. Continuous line: trend over the years.

**Table 5.** Results of ITS analysis of the monthly number of accesses, by demographic and socioeconomic variables and group of diagnoses.

| FMHEA characteristics | Mean (per month) | SD | Trend pre-COVID-19 | COVID-19 immediate effect | Trend post-COVID-19 |
| --- | --- | --- | --- | --- | --- |
|  |  |  | IRR (95%CI) | IRR (95%CI) | IRR (95%CI) |
| **Total** | 1182.5 | 269.2 | **0.992 (0.988-0.997)** | **0.636 (0.481-0.840)** | **1.018 (1.001-1.036)** |
| **Sex** |  |  |  |  |  |
| Males | 492.6 | 102.2 | **0.992 (0.988-0.997)** | **0.684 (0.534-0.878)** | **1.016 (1.001-1.032)** |
| Females | 690.0 | 170.5 | **0.993 (0.988-0.997)** | **0.602 (0.431-0.840)** | 1.020 (0.999-1.041) |
| **Age class** |  |  |  |  |  |
| ≤24 | 230.4 | 51.3 | 0.998 (0.994-1.003) | **0.559 (0.412-0.758)** | **1.029 (1.028-1.048)** |
| 25-34 | 146.5 | 42.6 | **0.988 (0.980-0.995)** | **0.637 (0.446-0.910)** | 1.019 (0.996-1.043) |
| 35-64 | 493.8 | 120.8 | **0.991 (0.985-0.996)** | **0.672 (0.506-0.893)** | 1.012 (0.994-1.030) |
| 65-74 | 115.2 | 23.6 | **0.993 (0.989-0.998)** | **0.697 (0.530-0.915)** | **1.021 (1.004-1.038)** |
| 75-84 | 134.3 | 31.0 | 0.994 (0.987-1.000) | **0.664 (0.499-0.885)** | **1.020 (1.003-1.037)** |
| 85+ | 62.3 | 20.3 | **0.993 (0.990-0.996)** | **0.502 (0.423-0.597)** | **1.015 (1.005-1.026)** |
| **Deprivation**<br>**level (quintiles)** |  |  |  |  |  |
| Low | 217.5 | 50.8 | **0.993 (0.987-0.999)** | **0.592 (0.444-0.790)** | **1.031 (1.012-1.049)** |
| Middle-low | 267.5 | 60.1 | **0.990 (0.986-0.995)** | **0.654 (0.502-0.851)** | **1.020 (1.004-1.037)** |
| Middle | 251.7 | 58.5 | **0.994 (0.989-0.998)** | **0.619 (0.461-0.832)** | 1.018 (0.999-1.037) |
| Middle- high | 178.8 | 43.9 | **0.993 (0.987-0.998)** | **0.659 (0.485-0.895)** | 1.013 (0.993-1.032) |
| High | 122.7 | 36.9 | **0.992 (0.984-0.999)** | 0.694 (0.470-1.027) | 0.999 (0.976-1.023) |
| **Citizenship** |  |  |  |  |  |
| Italians+HDCs | 1050.5 | 242.3 | **0.993 (0.989-0.997)** | **0.624 (0.473-0.824)** | **1.019 (1.001-1.037)** |
| Immigrants<br>from HMPCs | 117.9 | 24.3 | **0.991 (0.986-0.997)** | **0.736 (0.580-0.935)** | 1.013 (0.999-1.028) |
| **Group of**<br>**diagnoses** |  |  |  |  |  |
| Delirium/Mental<br>Confusion | 125.8 | 19.1 | 1.001 (0.998-1.005) | **0.811 (0.699-0.941)** | 1.006 (0.997-1.014) |
| Dependencies | 75.5 | 23.5 | 1.017 (0.990-1.044) | **0.605 (0.386-0.948)** | 1.011 (0.985-1.037) |
| Eating<br>Disorders | 3.4 | 2.3 | 1.018 (0.978-1.060) | 1.023 (0.517-2.027) | 1.008 (0.971-1.046) |
| Anxiety<br>Disorders | 520.2 | 147.4 | **0.991 (0.985-0.996)** | **0.658 (0.467-0.928)** | 1.007 (0.985-1.030) |
| Mood Disorders | 95.5 | 28.1 | **0.988 (0.982-0.994)** | **0.681 (0.516-0.898)** | 1.008 (0.991-1.026) |
| Personality Disorders | 9.2 | 5.7 | **0.947 (0.930-0.963)** | 1.186 (0.841-1.673) | 0.999 (0.989-1.009) |
| Dissociation | 3.8 | 1.9 | **0.979 (0.968-0.990)** | 0.894 (0.565-1.414) | 1.009 (0.975-1.043) |
| Unspecified Psychoses | 102.2 | 17.0 | 0.998 (0.993-1.002) | 0.916 (0.799-1.049) | 1.001 (0.991-1.010) |
| Adjustment Disorders | 14.2 | 5.0 | 0.995 (0.987-1.004) | **0.651 (0.472-0.898)** | 1.014 (0.996-1.032) |
| Post-Traumatic Stress and related Disorders | 25.4 | 8.2 | 0.997 (0.989-1.005) | **0.604 (0.411-0.890)** | 1.007 (0.981-1.035) |
| Somatization | 144.6 | 54.9 | 0.991 (0.983-1.000) | 0.389 (0.269-0.562) | 1.031 (1.009-1.054) |
| Schizophrenic Spectrum | 5.8 | 2.6 | **0.976 (0.972-0.980)** | **1.207 (1.071-1.360)** | **0.981 (0.971-0.990)** |

When FMHEAs were analysed based on the deprivation level, a significant immediate effect of COVID-19 was found for all the categories. Post-COVID-19, the monthly FMHEAs increased, but this increasing trend was significant only for those residing in less deprived areas (Fig 3).

Dashed line: counterfactual scenario. Horizontal dashed line: pre-pandemic level.

When we explored differences by sex, a similar trend between males and females emerged. Indeed, in both cases, monthly FMHEAs significantly dropped at the COVID pandemic outbreak, followed by a post-COVID increase (Supplementary Fig 1). The same trend was found by age class, although the highest immediate effect of COVID-19 was found on the youngest and the oldest subjects (IRR=0.559, 95%CI 0.412-0.758 and IRR=0.502, 95%CI 0.423-0.597, respectively) and when comparing the monthly number of FMHEAs by citizenship (Supplementary Fig 2-3).

Supplementary Fig 4 and 5 show the results of the ITS analysis for the monthly number of FMHEAs by group of psychiatric diagnoses. In the pre-COVID-19 period, a significant decreasing trend was found for Anxiety Disorders, Mood Disorders, Personality Disorders, Dissociation, and Schizophrenic Spectrum. A significant drop in FMHEAs at the outbreak of the pandemic was observed for Delirium/Mental Confusion (-19%), Dependencies, and Post-Traumatic Stress and related Disorders (both -40%), Adjustment and Anxiety Disorders (both -35%), and Mood Disorders (-32%). A countertendency, Schizophrenic Spectrum showed a significant increase in FMHEAs (+21%). When evaluating post-COVID-19 trends, most diagnostic groups showed an increasing but not significant trend, with the curve not reaching the counterfactual curve at the end of the observational period (except for Dissociation). A significant post-COVID-19 trend was observed for monthly FMHEAs for Schizophrenic Spectrum, which remained above the expected trend.

## Discussion

Our study investigated the effects of the COVID-19 pandemic on ED accesses related to a psychiatric condition in a large open cohort of NHS beneficiaries. Within this topic, we specifically focused on how COVID-19 differentially affected FMHEAs depending on deprivation level, citizenships, age, sex, and diagnosis.

Overall, we showed that COVID-19 reduced incidence rates of ED accesses by 29%. This effect was confirmed after adjusting for demographic and socioeconomic covariates. Monthly ED accesses dropped at the pandemic outbreak, and progressively increased thereafter, until exceeding the counterfactual line. We observed that COVID-19 increased pre-existing differences across deprivation levels by exerting a stronger detrimental effect on highly deprived individuals. Among age classes, the elderly were the most affected by COVID-19 in terms of increased risk of access. The pandemic did not modify pre-existing differences in FMHEA risk between sexes or citizenship. Our results are consistent with studies conducted in France and the UK [22,23], which observed a decrease in ED accesses at the pandemic onset, followed by an increase thereafter. A recent cross-country study on over 3 million ED accesses confirmed that a decrease had occurred in 2020 in highly developed countries [24]. On the other hand, other authors have shown an increase in ED accesses during COVID-19 [25, 26]. Several reasons may contribute to the observed findings on ED use in our sample, including a variation in mental health needs across the pandemic waves and concerns about using healthcare services due to the fear of infections. Regarding this latter aspect, Sanmarchi and collaborators [27] observed an overall decrease in ED use in Italy, where the strongest effect was observed in regions were COVID-19 hit the hardest, including Lombardy and Tuscany. It is noteworthy that the trend of monthly ED accesses exceeded the counterfactual line in June 2021, when weekly COVID cases were steadily decreasing, and 34 million people had already received at least the first dose of COVID-19 vaccine. Therefore, we can expect that patients were less concerned about contracting the infection when using healthcare services. On the other hand, it is unlikely that reduced ED accesses during 2020 could be attributable to better mental health.

Indeed, evidence from international studies points to an increase in self-reported depression and anxiety [28], and the prevalence and incidence of mental health conditions were also found to be slightly higher early in the pandemic [29].

We found inequalities in the risk of ED access for mental health conditions by deprivation level and citizenship.

Indeed, before COVID-19, the risk of FMHEAs went up as deprivation level increased. We can hypothesize that people living in more affluent areas used the ED for psychiatric emergencies less frequently either because they used other mental healthcare services or because they carried a lower psychiatric burden. The extent to which area deprivation affects mental health has been extensively documented [30]. Neighbourhood deprivation has been associated with poor mental health [31], and area deprivation explains regional variance in psychiatric morbidity [32]. However, whether increased psychiatric burden in deprived individuals results in a modification of mental healthcare service use is controversial. A lower socioeconomic index has been associated with increased community service contacts in psychiatric patients [33], whereas a lower mental health service use in the nonclinical population has been reported both in children and adolescents [34] and in the general population [35]. In studies specifically investigating ED use, deprivation has been suggested as a predisposing factor for access [36]. However, among specific socioeconomic factors, only unemployment could predict ED use for mental health conditions in the general population [37]. Differences in deprivation measures and social contexts may account for the observed discrepancies across studies.

The onset of the COVID-19 pandemic had a strong impact on the effects of deprivation on FMHEAs in that an inverse interactive effect was observed. Stronger effects of the pandemic were observed in people living in more deprived areas, so that pre-existing differences between deprivation levels were no longer observed after the pandemic outbreak. As the available evidence shows that poor socioeconomic conditions have been associated with an increased risk of developing psychiatric disorders after COVID-19 [38, 39, 40], an increased demand for psychiatric care would have been expected from highly deprived individuals. Whether and why these patients experienced limitations to ED use for psychiatric conditions when needed should be further investigated.

Immigrants from HMPCs showed a higher risk of ED access for psychiatric conditions than did Italians and immigrants from HDCs both before and after the COVID-19 outbreak. In both groups, rates of ED access decreased after the outbreak, and no interaction between COVID-19 and citizenship was detected. Evidence so far has shown that HMPCS immigrants are more likely to have psychiatric disorders [41], including because of the stress factors encountered during their migration path and their post-migration life conditions. As immigrants are less likely to use outpatient mental health services [42, 43, 44, 45, 46], the ED is the main health service where their mental health needs can be met. In addition, the lack of psychosocial support such as that provided by psychiatric outpatient services could facilitate relapse requiring urgent care.

FMHEA risk was higher in adolescents and young adults than in adults and the elderly, except those aged >85. Considering that most psychiatric disorders make their first appearance by an individual’s mid-20s [47], it is likely that increased ED referral is related to the acute onset of such conditions. COVID-19 hit the elderly (>64) stronger than it did other age classes, which is consistent with the increased vulnerability of older adults to the mental health effects of the pandemic [48] and their more challenging access to digital health [49], although, to our knowledge, this issue has not yet been extensively explored in the Italian context.

When we looked at the effect of sex, we observed that males had a lower risk of FMHEAs, and COVID-19 did not have any significant impact on this pattern. Sex differences in ED use for psychiatric conditions were found in previous studies, of which some pointed to an increased risk in females [50, 51], while others to a decrease [52,53]. This difference between sexes could be explained by the fact that females are more likely to acknowledge psychological distress with respect to males, but they mostly seek help from primary care services [54].

ITS analysis of monthly trends is particularly useful to investigating the differential effect of COVID-19 on the basis of the psychiatric diagnosis. Consistent with previous evidence, we found no significant variation in psychotic- or personality disorder-related ED accesses before and after the pandemic, while anxiety and mood disorders decreased at the pandemic outbreak [55, 56].

Regarding addiction-related FMHEAs, we found a drop after the outbreak, in contrast with another study that showed stable ED access across the pandemic waves [57]. Evidence so far points to an effect of COVID-19 on rising ED referrals for eating disorders in the pandemic phase [58], especially in the youngest age class [59], which we did not find in our data. However, few FMEHAs were related to eating disorders in our sample, which does not make it possible to draw any conclusion. It is interesting to note that trauma-related- FMHEAs (PTSD and Adjustment Disorders) significantly declined after the outbreak of the pandemic and did not change in the months following the pandemic outbreak. These results may seem surprising as COVID-19 is a potentially traumatic event because it involves exposure to or anticipation of life-threating experiences [60]. However, we can expect that the ED be viewed as a potential trigger by patients with COVID-related PTSD and that they may therefore avoid it. In our study, PTSD-related FMHEAs remained consistently well below the counterfactual trend. Regarding somatization, we observed a drop, followed by a steep and significant increase, which deserves further follow-up.

Indeed, an increased prevalence of somatic symptoms in infected individuals has been repeatedly documented, suggesting that somatization may be directly related to SARS-CoV2 infection [61, 62, 63, 64]. Also, it has been suggested that long COVID could represent a somatic syndrome [65]; indeed, having persistent COVID symptoms has been associated with more somatization [66].

The main strength of this study is its design, which allowed us to follow up a large cohort of about 6 million people, which represents nearly 10% of the Italian population. To our knowledge, this is the first study that longitudinally assessed the impact of COVID-19 on ED use for mental health conditions. Moreover, our period of observation covered the most crucial phases of the COVID-19 pandemic, from its outbreak, leading to mitigation measures, to the loosening of restrictions and post-vaccination.

Some limitations should be taken into account. First, we were not able to investigate long-term post-pandemic effects. However, the CoMeH study cohort was followed up until December 2024, thus allowing us to further explore the impact of COVID-19 on mental health beyond the pandemic once the data are available. Another limitation concerns potential residual ecological bias due to the use of an area-based deprivation index. Moreover, this approach could result in overlooking the role of individual socioeconomic markers such as income or education. Finally, the heterogeneity of the citizenship categories must be taken into account. Indeed, the “HMPCs immigrants” group includes both migrants that have obtained a residence permit and children of non-Italian citizens born and raised in Italy.

On the other hand, people born and raised abroad that have obtained Italian citizenship because of Italian ancestors have been included in the Italians group.

## Conclusion

Our study sheds light on inequalities in ED use related to psychiatric conditions before and after the COVID-19 outbreak. The mental health needs in more disadvantaged and vulnerable subjects need to be addressed in other healthcare settings, as the ED should be primarily dedicated to providing immediate care for severe health conditions. The transition of mental healthcare from the ED to the community must be encouraged, tailored on the basis of individual needs, and involve cultural-linguistic mediators, social workers, and/or outreach activities. At the research level, follow-up studies are recommended in order to unravel post-pandemic trajectories of mental disorders.

## Data Availability

All relevant data are within the manuscript and its Supporting Information files.

S1Fig: Number of monthly FMHSAs before and during the COVID-19 pandemic by sex. Vertical dashed line: introduction of restrictions. Continuous line: trend over the years. Dashed line: counterfactual scenario. Horizontal dashed line: pre-pandemic level.

S2Fig: Number of monthly FMHSAs before and during the COVID-19 pandemic by age. Vertical dashed line: introduction of restrictions. Continuous line: trend over the years. Dashed line: counterfactual scenario. Horizontal dashed line: pre-pandemic level.

S3Fig: Number of monthly FMHSAs before and during the COVID-19 pandemic by citizenship. Vertical dashed line: introduction of restrictions. Continuous line: trend over the years. Dashed line: counterfactual scenario. Horizontal dashed line: pre-pandemic level.

S4Fig: Number of monthly FMHSAs before and during the COVID-19 pandemic for Delirium, Eating Disorders, Unspecified Psychoses, Dependencies, Personality and Schizophrenic Disorders. Vertical dashed line: introduction of restrictions. Continuous line: trend over the years. Dashed line: counterfactual scenario. Horizontal dashed line: pre-pandemic level.

S5Fig: Number of monthly FMHSAs before and during the COVID-19 pandemic for Anxiety Disorders, PTSD, Adjustment Disorders, Mood Disorders, Somatization, and Dissociation. Vertical dashed line: introduction of restrictions. Continuous line: trend over the years. Dashed line: counterfactual scenario. Horizontal dashed line: pre-pandemic level.

## Acknowledgements

We thank Jacqueline M. Costa for her contribution in translating and copy editing the manuscript.

## References

1. Nicola M, Alsafi Z, Sohrabi C, Kerwan A, Al-Jabir A, Iosifidis C et al. The socio-economic implications of the coronavirus pandemic (COVID-19): A review. Int J Surg. 2020 Jun;78:185–193

2. Liu S, Haucke MN, Heinzel S, Heinz A. Long-Term Impact of Economic Downturn and Loneliness on Psychological Distress: Triple Crises of COVID-19 Pandemic. Journal of Clinical Medicine. 2021; 10(19):4596

3. Rondinelli C, Zanichelli F. Principali risultati della terza edizione dell’indagine straordinaria sule famiglie italiane 2020.Note COVID-19. 30 marzo 2021.Banca d Italia.

4. Bonnini S, Borghesi M (2022). Relationship between Mental Health and SocioEconomic, Demographic and Environmental factors in the COVID-19 Lockdown Period - A multivariate Regression Analysis .Mathematics, MDPI, vol 10 (18),Pages 1-15,September

5. Robinson E, Sutin AR, Daly M, Jones A. A systematic review and meta-analysis of longitudinal cohort studies comparing mental health before versus during the COVID-19 pandemic in 2020. J Affect Disord. 2022 Jan 1;296:567–576.

6. World Health organization. Mental Health and COVID-19: Early Evidence of the Pandemic’s Impact: Scientific Brief, 2 March 2022 (who.int). Available online: https://www.who.int/publications-detail-redirect/WHO-2019-nCoV-Sci_Brief-Mental_health-2022.1.

7. Wu T, Jia X, Shi H, Niu J, Yin X, Xie J, Wang X. Prevalence of mental health problems during the COVID-19 pandemic: A systematic review and meta-analysis. J Affect Disord. 2021 Feb 15;281:91–98

8. Parenteau AM, Boyer CJ, Campos LJ, Carranza AF, Deer LK, Hartman DT et al. A review of mental health disparities during COVID-19: Evidence, mechanisms, and policy recommendations for promoting societal resilience. Dev Psychopathol. 2023 Oct;35(4):1821–1842

9. Sun Y, Wu Y, Fan S, Dal Santo T, Li L, Jiang X et al.. Comparison of mental health symptoms before and during the covid-19 pandemic: evidence from a systematic review and meta-analysis of 134 cohorts. BMJ. 2023 Mar 8;380:e074224.

10. Aragona M, Tumiati MC, Ferrari F, VialeS, Nicolella G, Barbato A et al. Psychopathological effects of the Coronavirus (Sars-CoV-2) imposed Lockdown on vulnerable patients in treatment in a Mental Health outpatient department for migrants and individuals in poor SocioEconomic conditions. Int J Soc Psychiatry. 2022 Feb; 68(1):203–209

11. Duden GS, Gersdorf S, Stengler K. Global impact of the COVID-19 pandemic on mental health services: A systematic review. J Psychiatr Res. 2022 Oct;154:354–377

12. Rojnic Kuzman M, Vahip S, Fiorillo A, Beezhold J, Pinto da Costa M, Skugarevsky O et al. Mental health services during the first wave of the COVID-19 pandemic in Europe: Results from the EPA Ambassadors Survey and implications for clinical practice. Eur Psychiatry. 2021 Jun 9;64(1):e41.

13. Carpiniello B, Tusconi M, Zanalda E, Di Sciascio G, Di Giannantonio M . Executive Committee of The Italian Society of Psychiatry. Psychiatry during the Covid-19 pandemic: a survey on mental health departments in Italy. BMC Psychiatry. 2020 Dec 16;20(1):593.

14. Di Girolamo C, Gnavi R, Landriscina T, Forni S, Falcone M, Calandrini E et al.Indirect impact of the COVID-19 pandemic and its containment measures on social inequalities in hospital utilization in Italy.J Epidemiol Community Health. 2022 May12:jech-2021-218452.

15. Petrelli A, Aragona M, Ciampichini R, Di Napoli A, Fano V, Leone S et al. A population-based cohort to investigate the impact of COVID-19 on socioeconomic inequalities in mental health care in Italy (CoMeH): study protocol. Soc Psychiatry Psychiatr Epidemiol. 2025 Feb 14. doi: 10.1007/s00127-025-02838-y.

16. Capuzzi E, Di Brita C, Caldiroli A, Colmegna F, Nava R, Buoli M et al. Psychiatric emergency care during Coronavirus 2019 (COVID 19) pandemic lockdown: results from a Department of Mental Health and Addiction of northern Italy. Psychiatry Res. 2020 Nov;293:113463..

17. Turcato G, Zaboli A, Luchetti A, Sighele F, Sibilio S, Donato C et al. "Effect of the SARS-COV-2 pandemic outbreak on the emergency department admission for an acute psychiatric condition". J Psychiatr Res. 2022 Jul;151:626–632.

18. Petrelli A, Aragona M, Ciampichini R, Di Napoli A, Fano V, Leone S et al. A population-based cohort to investigate the impact of COVID-19 on socioeconomic inequalities in mental health care in Italy (CoMeH): study protocol. Soc Psychiatry Psychiatr Epidemiol. 2025 Feb 14. doi: 10.1007/s00127-025-02838-y. Epub ahead of print. PMID: 39953166.

19. Rosano A, Pacelli B, Zengarini N, Costa G, Cislaghi C, Caranci N. Aggiornamento e revisione dell’indice di deprivazione italiano 2011 a livello di sezione di censimento [Update and review of the 2011 Italian deprivation index calculated at the census section level]. Epidemiol Prev. 2020 Mar-Jun;44(2-3):162-170

20. Pacelli B, Zengarini N, Broccoli S, Caranci N, Spadea T, Di Girolamo C, et al. Differences in mortality by immigrant status in Italy. Results of the Italian Network of Longitudinal Metropolitan Studies. Eur J Epidemiol. (2016) 31:691–701

21. Schuengel C, Tummers J, Embregts P, Leusink GL. Impact of the initial response to COVID-19 on long-term care for people with intellectual disability: an interrupted time series analysis of incident reports. J Intellect Disabil Res. (2020) 64:817–24.

22. Akkaoui MA, Barruel D, Dauriac-Le Masson V, Gourevitch R, Pham-Scottez A. Trends in psychiatric emergency visits: insights from France’s largest psychiatric emergency department. Int J Emerg Med. 2025 Jan 15;18(1):13

23. Jacob N, Santos R, Sivey P. The long-run effect of COVID-19 on hospital emergency department attendances:evidence from statistical analysis of hospital data from England. Health Policy. 2024 Dec;150:105168. Erratum in: Health Policy. 2024 Nov 29;152:105217.

24. Bowden N, Hedquist A, Dai D, Abiona O, Bernal-Delgado E, Blankart CR et al. International comparison of hospitalizations and emergency department visits related to mental health conditions across high-income countries before and during the COVID-19 pandemic. Health Serv Res. 2024 Dec;59(6):e14386

25. Aymerich C, Pedruzo B, Salazar de Pablo G, Olazabal N, Catalan A, González-Torres MÁ. Number and nature of psychiatric emergency department visits in a tertiary hospital before, during, and after coronavirus pandemic. Front Psychiatry. 2024 Apr 18;15:1380401..

26. Mustafa FA. No reduction in psychiatric emergency department visits during COVID-19: A downside of telepsychiatry? Int J Soc Psychiatry. 2021 Jun;67(4):402–403.

27. Sanmarchi F, Golinelli D, Lenzi J, Grilli R, Fantini MP, Gori D. The impact of the COVID- 19 pandemic on the use of Emergency Departments in Italy. Ann Ig. 2023 Jul-Aug;35(4):413-424

28. Pennix BWJH, Benros ME, Klein RS, Vinkers CH. How COVID-19 shaped Mental Health: from infection to pandemic effects. Nat.Med.2022 Oct; 28(10):2027–2037.

29. Ahmed N, Barnett P, Greenburgh A, Pemovska T, Stefanidou T, Lyons N et al. Mental health in Europe during the COVID-19 pandemic: a systematic review. Lancet Psychiatry. 2023 Jul;10(7):537–556.

30. Kivimäki M, Batty GD, Pentti J, Shipley MJ, Sipilä PN, Nyberg ST et al. Association between socioeconomic status and the development of mental and physical health conditions in adulthood: a multi-cohort study. Lancet Public Health. 2020 Mar;5(3):e140–e149.

31. Fone D, White J, Farewell D, Kelly M, John G, Lloyd K et al. Effect of neighbourhood deprivation and social cohesion on mental health inequality: a multilevel population-based longitudinal study. Psychol Med. 2014 Aug;44(11):2449–60.

32. Skapinakis P, Lewis G, Araya R, Jones K, Williams G. Mental health inequalities in Wales, UK: multi-level investigation of the effect of area deprivation. Br J Psychiatry. 2005 May;186:417-22.

33. Donisi V, Tedeschi F, Percudani M, Fiorillo A, Confalonieri L, De Rosa C et al. Prediction of community mental health service utilization by individual and ecological level socio- economic factors. Psychiatry Res. 2013 Oct 30;209(3):691–8.

34. Sharifi V, Dimitropoulos G, Williams JVA, Rao S, Pedram P, Bulloch AGM, Patten SB. Neighborhood material versus social deprivation in Canada: different patterns of associations with child and adolescent mental health problems. Soc Psychiatry Psychiatr Epidemiol. 2024 May

35. Ngamini Ngui A, Perreault M, Fleury MJ, Caron J. A multi-level study of the determinants of mental health service utilization. Rev Epidemiol Sante Publique. 2012 Apr;60(2):85–93.

36. Fleury MJ, Rochette L, Grenier G, Huỳnh C, Vasiliadis HM, Pelletier É, Lesage A. Factors associated with emergency department use for mental health reasons among low, moderate and high users. Gen Hosp Psychiatry. 2019 Sep-Oct;60:111-119.

37. Saini P, McIntyre J, Corcoran R, Daras K, Giebel C, Fuller E et al. Predictors of emergency department and GP use among patients with mental health conditions: a public health survey. Br J Gen Pract. 2019 Dec 26;70(690):e1–e8. 0.

38. Hubbard G, den Daas C, Johnston M, Dixon D. Sociodemographic and Psychological Risk Factors for Anxiety and Depression: Findings from the Covid-19 Health and Adherence Research in Scotland on Mental Health (CHARIS-MH) Cross-sectional Survey. Int J Behav Med. 2021 Dec;28(6):788–800

39. Camara C, Surkan PJ, Van Der Waerden J, Tortelli A, Downes N, Vuillermoz C et al. COVID-19-related mental health difficulties among marginalised populations: A literature review. Glob Ment Health (Camb). 2022 Dec 9;10:e2

40. Kim J, Linos E, Rodriguez CI, Chen ML, Dove MS, Keegan TH. Prevalence and associations of poor mental health in the third year of COVID-19: U.S. population-based analysis from 2020 to 2022. Psychiatry Res. 2023 Dec;330:115622

41. Priebe S, Giacco D, El-Nagib R. Public Health Aspects of Mental Health Among Migrants and Refugees: A Review of the Evidence on Mental Health Care for Refugees, Asylum Seekers and Irregular Migrants in the WHO European Region [Internet]. Copenhagen: WHO Regional Office for Europe; 2016. (Health Evidence Network Synthesis Report, No. 47.) Available from: https://www.ncbi.nlm.nih.gov/books/NBK391045/

42. Gubi E, Sjöqvist H, Viksten-Assel K, Bäärnhielm S, Dalman C, Hollander AC. Mental health service use among migrant and Swedish-born children and youth: a register-based cohort study of 472,129 individuals in Stockholm. Soc Psychiatry Psychiatr Epidemiol. 2022 Jan;57(1):161–171.

43. Tortelli A, Perquier F, Melchior M et al (2020). Mental Health and Service Use of Migrants in Contact with the Public Psychiatry System in Paris. Int J Environ Res Public Health. Dec 15;17(24):9397

44. Cristofalo D, Bonetto C, Ballarin M, Amaddeo F, Ruggeri M, Nosè M et al. Access to and Use of Psychiatric Services by Migrants Resettled in Northern Italy. J Immigr Minor Health. 2018 Dec;20(6):1309–1316.

45. Aragona M, Tumiati MC, Ferrari F, VialeS, Nicolella G, Barbato A et al. Psychopathological effects of the Coronavirus (Sars-CoV-2) imposed Lockdown on vulnerable patients in treatment in a Mental Health outpatient department for migrants and individuals in poor SocioEconomic conditions. Int J Soc Psychiatry. 2022 Feb; 68(1):203–209.

46. Benjamen J, Girard V, Jamani S, Magwood O, Holland T, Sharfuddin N rt al. Access to Refugee and Migrant Mental Health Care Services during the First Six Months of the COVID-19 Pandemic: A Canadian Refugee Clinician Survey. Int J Environ Res Public Health. 2021 May 15;18(10):5266

47. Solmi M, Radua J, Olivola M, Croce E, Soardo L, Salazar de Pablo G et al. Age at onset of mental disorders worldwide: large-scale meta-analysis of 192 epidemiological studies. Mol Psychiatry. 2022 Jan;27(1):281–295.

48. Lau J, Koh WL, Ng JS, Khoo AM, Tan KK. Understanding the mental health impact of COVID-19 in the elderly general population: A scoping review of global literature from the first year of the pandemic. Psychiatry Res. 2023 Nov;329:115516.

49. Vakkalanka JP, Gadag K, Lavin L, Ternes S, Healy HS, Merchant KAS et al. Telehealth Use and Health Equity for Mental Health and Substance Use Disorder During the COVID- 19 Pandemic: A Systematic Review. Telemed J E Health. 2024 May;30(5):1205–1220.

50. Lucero AD, Lee A, Hyun J, Lee C, Kahwaji C, Miller G et al. Underutilization of the Emergency Department During the COVID-19 Pandemic. West J Emerg Med. 2020 Sep 24;21(6):15–23.

51. Perozziello A, Sousa D, Aubriot B, Dauriac-Le Masson V. Use of mental health services in the aftermath of COVID-19 waves: a retrospective study conducted in a French Psychiatric and Neurosciences University Hospital. BMJ Open. 2023 Feb 23;13(2):e064305.

52. Beghi M, Brandolini R, Casolaro I, Beghi E, Cornaggia CM, Fraticelli C et al. Effects of lockdown on emergency room admissions for psychiatric evaluation: an observational study from the AUSL Romagna, Italy. Int J Psychiatry Clin Pract. 2021 Jun;25(2):135–139

53. Lavergne MR, Shirmaleki M, Loyal JP, Jones W, Nicholls TL, Schütz CG et al. Emergency department use for mental and substance use disorders: descriptive analysis of population-based, linked administrative data in British Columbia, Canada. BMJ Open. 2022 Jan 13;12(1):e057072.

54. World Health organization. Mental Health and COVID-19: Early Evidence of the Pandemic’s Impact: Scientific Brief, 2 March 2022 (who.int). Available online: https://www.who.int/publications-detail-redirect/WHO-2019-nCoV-Sci_Brief-Mental_health-2022.1.

55. Gonçalves-Pinho M, Mota P, Ribeiro J, Macedo S, Freitas A. The Impact of COVID-19 Pandemic on Psychiatric Emergency Department Visits - A Descriptive Study. Psychiatr Q. 2021 Jun;92(2):621–631.

56. Hamlin M, Ymerson T, Carlsen HK, Dellepiane M, Falk Ö, Ioannou M et al. Changes in Psychiatric Inpatient Service Utilization During the First and Second Waves of the COVID- 19 Pandemic. Front Psychiatry. 2022 Feb 17;13:829374.

57. Håkansson A, Grudet C. Decreasing Psychiatric Emergency Visits, but Stable Addiction Emergency Visits, During COVID-19-A Time Series Analysis 10 Months Into the Pandemic. Front Psychiatry. 2021 Jul 13;12:664204.

58. Castellini G, Cassioli E, Rossi E . Use and misuse of the emergency room by patients with eating disorders in a matched-cohort analysis: What can we learn from it? Psychiatry Res. 2023 Oct;328:115427.

59. Toulany A, Saunders NR, Kurdyak P, Strauss R, Fu L, Joh-Carnella N et al. Acute presentations of eating disorders among adolescents and adults before and during the COVID-19 pandemic in Ontario, Canada. CMAJ. 2023 Oct 3;195(38):E1291–E1299.

60. Bridgland VME, Moeck EK, Green DM, Swain TL, Nayda DM, Matson LA et al. Why the COVID-19 pandemic is a traumatic stressor. PLoS One. 2021 Jan 11;16(1):e0240146.

61. Tröscher A, Gebetsroither P, Rindler M, Böhm V, Dormann R, von Oertzen T, et al. High Somatization Rates, Frequent Spontaneous Recovery, and a Lack of Organic Biomarkers in Post-Covid-19 Condition. Brain Behav. 2024 Oct;14(10):e70087

62. Lier J, Stoll K, Obrig H, Baum P, Deterding L, Bernsdorff N et al. Neuropsychiatric phenotype of post COVID-19 syndrome in non-hospitalized patients. Front Neurol. 2022 Sep 27;13:988359

63. Dong F, Liu HL, Dai N, Yang M, Liu JP. A living systematic review of the psychological problems in people suffering from COVID-19. J Affect Disord. 2021 Sep 1;292:172–188.

64. Noviello D, Costantino A, Muscatello A, Bandera A, Consonni D, Vecchi M et al. Functional gastrointestinal and somatoform symptoms five months after SARS-CoV-2 infection: A controlled cohort study. Neurogastroenterol Motil. 2022 Feb;34(2):e14187

65. Joffe AR, Elliott A. Long COVID as a functional somatic symptom disorder caused by abnormally precise prior expectations during Bayesian perceptual processing: A new hypothesis and implications for pandemic response. SAGE Open Med. 2023 Aug 24;11:20503121231194400

66. Nehme A, Barakat M, Malaeb D, Obeid S, Hallit S, Haddad G. Association between COVID-19 symptoms, COVID-19 vaccine, and somatization among a sample of the Lebanese adults. Pharm Pract (Granada). 2023 Jan-Mar;21(1):2763

